# The Mendelian randomization analysis of trace elements Zinc, Selenium, Copper, Iron, and Phosphate with congenital foot deformities

**DOI:** 10.1101/2024.10.26.24316184

**Authors:** Ling Long, Jiaxiang Gu, Haifeng You, Xiaobing Xiang, Wenyu Wu, Gaorong Deng

**Affiliations:** Department of orthopaedics, Jiujiang Hospital of Traditional Chinese Medicine, 555 Dehua Road, Jiujiang, 332000, Jiangxi, China; Department of Pediatric Orthopaedics, Beijing Jishuitan Hospital, Capital Medical University (National Orthopaedic Medical Center), No. 31, Xinjiekou East Street, Xicheng District, Beijing, 100035, Beijing, China; Jiangxi University of Chinese Medicine, Organization, 1688 Meiling Avenue,Wanli District, Nanchang, 330000, Nanchang, China; Nanchang University Affiliated Rehabilitation Hospital, Orthopedics Department, No. 133, South Square Road, Nanchang, 330000, Nanchang, China

**Author notes:** Contributing authors.

**Keywords:** Mendelian randomization, trace elements, Zinc, Selenium, Copper, Iron, Phosphate, congenital foot deformities, gut microbiota

## Abstract

Congenital foot deformities are complex diseases influenced by genetic, environmental, and nutritional factors. In recent years, research has increasingly focused on the effects of trace elements during pregnancy (such as zinc, selenium, copper, iron, and phosphate) on fetal development. Although observational studies suggest that these trace elements may be associated with congenital deformities, their causal relationship remains unclear. This study employs the Mendelian Randomization method, using genetic variants as instrumental variables for analysis. The results show that although the OR for Zinc is 92.21 (95% CI: 0.0008–10098024, *p* = 0.445), for Selenium the OR is 6.21 × 10^-6^ (95% CI: 2.71 × 10^-15^–14233.68, *p* = 0.276), for Copper the OR is 0.96 (95% CI: 0.823–1.128, *p* = 0.646), for Iron the OR is 0.023 (95% CI: 2.10 × 10^-8^–24289.32, *p* = 0.593), and for Phosphate the OR is 0.96 (95% CI: 0.667–1.374, *p* = 0.812). These data did not reach statistical significance, suggesting that the causal relationship between the levels of these trace elements and congenital foot deformities is unclear. However, further analysis revealed that selenium may reduce the risk of congenital foot deformities by regulating the gut microbiota class.Bacilli, with a significant interaction observed between selenium and this microbiota. Although sensitivity analyses (IVW and MR Egger methods) were unable to confirm the direct effect of selenium on congenital foot deformities, considering the role of class.Bacilli.id.1673 strengthened selenium’s indirect effect. Despite the overall results not reaching statistical significance, this study provides new clues for further exploration of the complex relationships between trace elements, gut microbiota, and congenital deformities, particularly the potential role of gut microbiota in modulating the relationship between nutritional elements and developmental abnormalities.

## 1 Introduction

In recent years, research has gradually revealed the important roles of trace elements such as Zinc, Selenium, Copper, Iron, and Phosphate in embryonic development[1]. These elements play indispensable roles in critical biological processes, including cell growth, immune function, oxidative stress regulation, and bone mineralization[2]. Deficiency or metabolic abnormalities of these elements may lead to a series of complex physiological disorders, potentially resulting in congenital foot deformities[3], such as clubfoot[4]. Zinc is involved in DNA synthesis and cell proliferation and is essential for the normal development of fetal organs[5]. Selenium, with its antioxidant properties, protects cells from oxidative stress damage, reducing the risk of fetal developmental abnormalities[6]. Copper is crucial for collagen synthesis, nervous system development, and iron metabolism, and its deficiency may lead to connective tissue malformations, thereby causing skeletal deformities[7]. Iron is not only a core component of hemoglobin but also plays a role in oxygen transport and energy metabolism. Iron deficiency may result in fetal hypoxia, which can affect the normal development of bones and muscles[8]. Phosphate is a vital component of bone structure, playing a key role in bone mineralization, and any metabolic imbalance can lead to abnormal skeletal development[9]. There may be complex interactions among these trace elements that affect various aspects of fetal development[10]. Although their importance in embryonic development is widely recognized, the causal relationship between these elements and congenital foot deformities remains unclear, warranting further research to provide more definitive scientific evidence.

Trace elements such as Zinc, Selenium, Copper, Iron, and Phosphate exhibit complex interactions during fetal development, playing a crucial role in maintaining normal physiological functions and promoting healthy development[11]. Firstly, the metabolism of Copper and Iron is closely related, with Copper being an essential cofactor in Iron metabolism, involved in the absorption of Iron from the intestines, its transport, and its utilization by cells[12]. A deficiency in Copper can lead to reduced effective utilization of Iron, resulting in anemia, which affects the oxygen supply to the fetus and overall development[13], particularly in the development of bones and muscles[14]. Secondly, Zinc and Phosphate have a synergistic effect on bone development[15]. A deficiency in Zinc may lead to abnormal Phosphate metabolism, thereby affecting bone mineralization and increasing the risk of skeletal developmental abnormalities[16]. Additionally, Selenium, through its powerful antioxidant functions, can reduce oxidative stress damage to cells, with oxidative stress being a major factor leading to Iron metabolism disorders[17]. Adequate intake of Selenium can mitigate the negative effects of Iron metabolism disorders, indirectly protecting fetal bone development and preventing skeletal developmental disorders caused by Iron metabolism imbalances[18]. Overall, these trace elements interact in a complex network during embryonic development, collectively supporting the normal developmental process. A deficiency or metabolic abnormality in any of these elements can have profound effects on the healthy development of the fetus[19].

This study employs the Mendelian randomization method to investigate the causal relationship between the minerals and dietary supplements Zinc, Selenium, Copper, Iron, and Phosphate and the risk of congenital foot deformities[20]. Mendelian randomization is an analytical technique that uses genes as instrumental variables, effectively eliminating confounding factors in observational studies, thereby allowing for more accurate inferences about the causal relationship between exposure factors and health outcomes[21]. We hypothesize that these trace elements may have a causal association with the occurrence of congenital foot deformities[22]. To test this hypothesis, we conducted a systematic evaluation of the relationship between these trace elements and congenital foot deformities using Mendelian randomization analysis, and we applied sensitivity analysis methods such as the Inverse Variance Weighted (IVW) method[23] and MR-Egger regression[24]to ensure the robustness of the results. The significance of this study lies in its potential to provide new genetic evidence for understanding the role of these trace elements in fetal development[25], as well as to offer a theoretical basis for nutritional management during pregnancy, helping to prevent the occurrence of congenital foot deformities[26]. We hope that this research will fill gaps in existing studies, provide references for future research and clinical practice, and offer scientific support for optimizing trace element intake during pregnancy to reduce the risk of congenital deformities.

## 2 Results

### 2.1 The causal relationship between Zinc and congenital foot deformities

The results of the Inverse Variance Weighted (IVW) analysis (Figure1) indicate that the causal association between Zinc levels and congenital foot deformities did not reach statistical significance (OR = 92.21, 95% CI: 0.000842–10098024, *p* = 0.4448) (Table 6). Although the OR value is relatively large, suggesting a potential influence of Zinc, the extremely wide confidence interval reflects a high degree of uncertainty in the results. The MR Egger regression analysis shows an OR value for Zinc of 1282875251 (95% CI: 8.925–1.84394*×*10^17^, *p* = 0.0475) (Table3), where the *p*-value reaches statistical significance. However, the extremely broad confidence interval suggests considerable uncertainty and potential bias. Additionally, the intercept of the Egger regression did not reach significance (*p* = 0.0561), implying that pleiotropy’s effect on the results may be limited.

**Table 1.** Data Sources for Exposure Factors.

| trace element | Data source | Sample size | Number of cases | Number of controls | Number of SNPs | Population | Data year |
| --- | --- | --- | --- | --- | --- | --- | --- |
| Zinc | UK Biobank | 336,314 | 13,151 | 323,163 | 10,894,596 | European population | 2017 |
| Selenium | UK Biobank | 461,384 | 11,059 | 450,325 | 9,851,867 | European population | 2018 |
| Copper | UK Biobank | 2,603 | NA | NA | 2,543,646 | European population | 2013 |
| Iron | UK Biobank | 461,384 | 14,150 | 447,234 | 9,851,867 | European population | 2018 |
| Phosphate | UK Biobank | 7,685 | 7,685 | NA | 9,811,692 | South Asian population | 2020 |

**Table 2.**
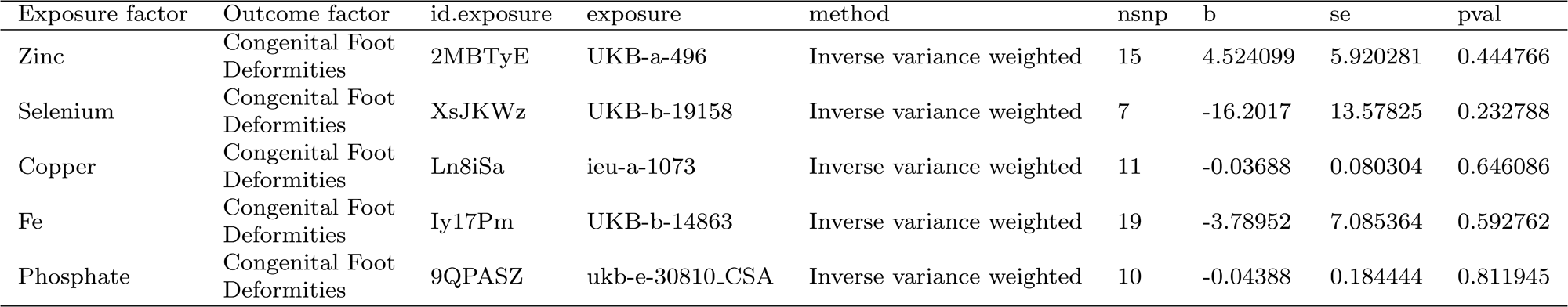
Inverse Variance Weighted (IVW) Method Results Data.

**Table 3.**
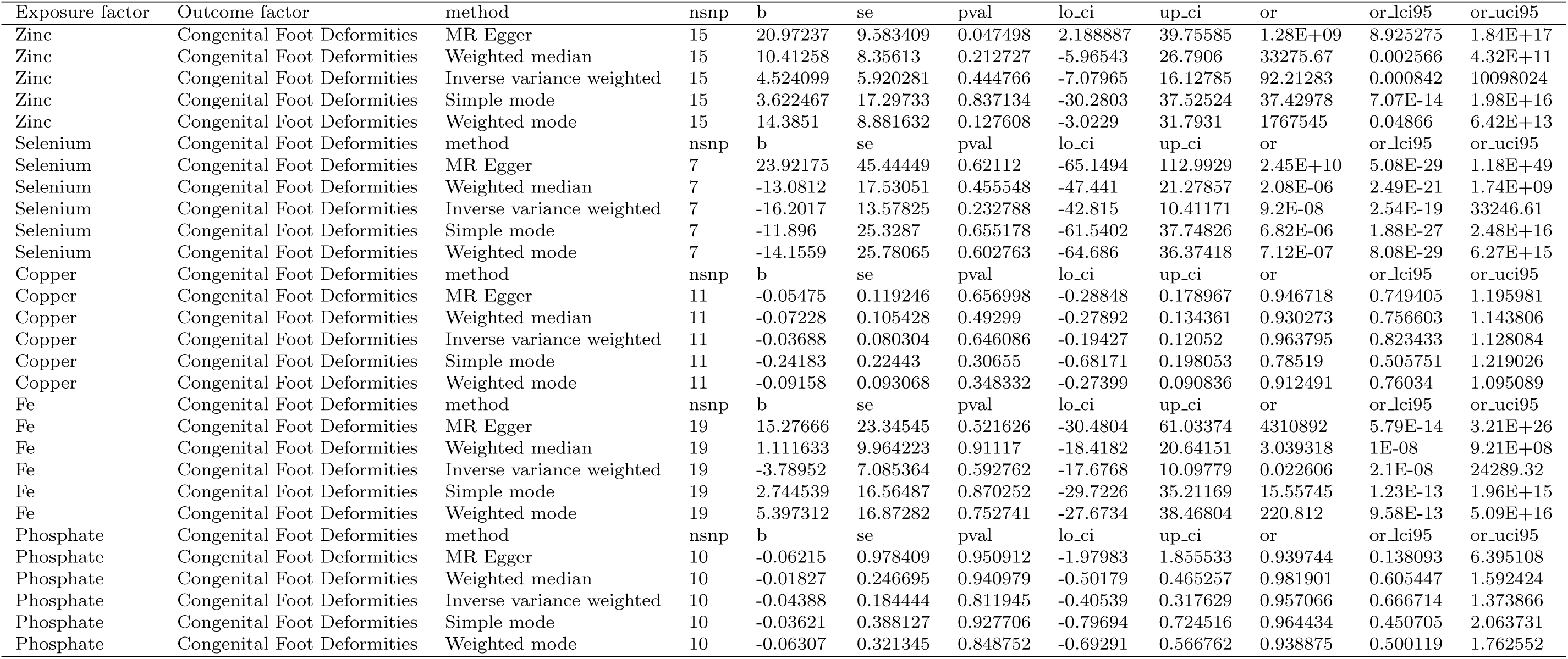
The Mendelian Randomization 5 Methods for trace element and congenital foot deformities.

**Table 4.**
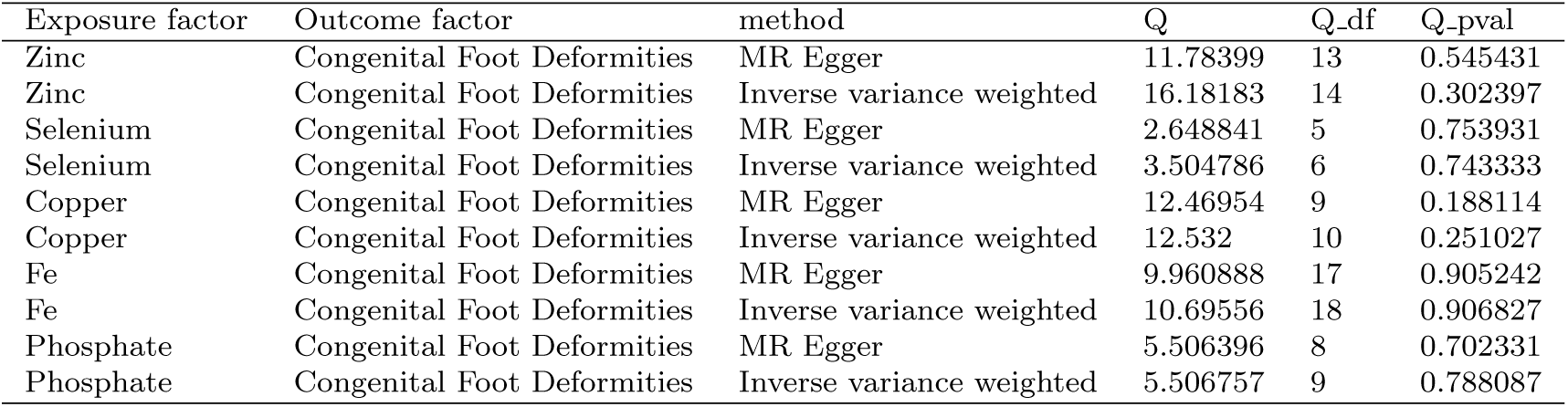
Heterogeneity Test of Exposure Factors.

| Exposure factor | Outcome factor | method | Q | Q_df | Q_pval |
| --- | --- | --- | --- | --- | --- |
| Zinc | Congenital Foot Deformities | MR Egger | 11.78399 | 13 | 0.545431 |
| Zinc | Congenital Foot Deformities | Inverse variance weighted | 16.18183 | 14 | 0.302397 |
| Selenium | Congenital Foot Deformities | MR Egger | 2.648841 | 5 | 0.753931 |
| Selenium | Congenital Foot Deformities | Inverse variance weighted | 3.504786 | 6 | 0.743333 |
| Copper | Congenital Foot Deformities | MR Egger | 12.46954 | 9 | 0.188114 |
| Copper | Congenital Foot Deformities | Inverse variance weighted | 12.532 | 10 | 0.251027 |
| Fe | Congenital Foot Deformities | MR Egger | 9.960888 | 17 | 0.905242 |
| Fe | Congenital Foot Deformities | Inverse variance weighted | 10.69556 | 18 | 0.906827 |
| Phosphate | Congenital Foot Deformities | MR Egger | 5.506396 | 8 | 0.702331 |
| Phosphate | Congenital Foot Deformities | Inverse variance weighted | 5.506757 | 9 | 0.788087 |

**Table 5.** Pleiotropy Test of Exposure factors Levels.

| Exposure factor | Outcome factor | exposure | egger_intercept | se | pval |
| --- | --- | --- | --- | --- | --- |
| Zinc | Congenital Foot Deformities | UKB-a-496 | -0.07522 | 0.03587 | 0.056107 |
| Selenium | Congenital Foot Deformities | UKB-b-19158 | -0.0812 | 0.087768 | 0.397333 |
| Copper | Congenital Foot Deformities | ieu-a-1073 | 0.007008 | 0.033005 | 0.836584 |
| Fe | Congenital Foot Deformities | UKB-b-14863 | -0.05002 | 0.058353 | 0.403296 |
| Phosphate | Congenital Foot Deformities | ukb-e-30810.CSA | 0.002635 | 0.138639 | 0.9853 |

**Table 6.** Odds Ratio (OR) and Confidence Interval (CI) Data.

| Exposure factor | Outcome factor | p_value | OR | CI_lower | CI_upper |
| --- | --- | --- | --- | --- | --- |
| Zinc | Congenital Foot Deformities | 0.444766 | 92.21283 | 0.000842 | 10098024 |
| Selenium | Congenital Foot Deformities | 0.232788 | 9.2E-08 | 2.54E-19 | 33246.61 |
| Copper | Congenital Foot Deformities | 0.646086 | 0.963795 | 0.823433 | 1.128084 |
| Fe | Congenital Foot Deformities | 0.592762 | 0.022606 | 2.1E-08 | 24289.32 |
| Phosphate | Congenital Foot Deformities | 0.811945 | 0.957066 | 0.666714 | 1.373866 |

The results of the Weighted Median method and the Simple Mode method also did not show significance (Table 3), further supporting the unclear causal relationship between Zinc and congenital foot deformities. The heterogeneity tests indicated that the Q statistics for both the IVW and MR Egger methods were not significant (Table 4), suggesting a high degree of consistency among the effects of the SNPs, with no significant heterogeneity observed. Additionally, the pleiotropy test showed that the p-value for the Egger intercept was close to significance but did not reach statistical significance (Table 5), implying that the influence of pleiotropy might be small but cannot be entirely ruled out.

These analysis results indicate that, although the IVW and MR Egger methods suggest a possible association between Zinc and congenital foot deformities (Tables 3 and 4), the wide confidence intervals and lack of statistical significance for the p-values imply substantial uncertainty and potential bias in the study findings. This suggests that the current data are insufficient to provide strong evidence for a clear causal relationship between Zinc and congenital foot deformities. Particularly, while the MR Egger regression shows statistical significance, the possible influence of unrecognized pleiotropy weakens the reliability of the conclusions. Although the Egger intercept in the pleiotropy test is close to significance (p = 0.056) (Table 3), it does not reach statistical significance.

### 2.2 The causal relationship between Selenium and congenital foot deformities

Using Mendelian randomization, we systematically evaluated the causal relationship between Selenium as an exposure factor and the risk of congenital foot deformities. The Inverse Variance Weighted (IVW) analysis results indicated that Selenium levels did not show a significant causal association with congenital foot deformities (OR = 9.1983*×*10^-8^, 95% CI: 2.54488*×*10^-19^–33246.61043, *p* = 0.2328) (Table 6). Although the OR value is extremely small, the vast range of the confidence interval suggests a high degree of uncertainty, possibly indicating that Selenium has a weak or negligible effect on congenital foot deformities. Similarly, the MR Egger regression analysis showed a higher OR value for Selenium (OR = 2.4495*×*10^10^, 95% CI: 5.08111*×*10^-29^– 1.1809*×*10^49^, *p* = 0.6211) (Table 3). However, the extremely wide confidence interval and the lack of statistical significance in the *p*-value further reflect the uncertainty of the causal association between Selenium and congenital foot deformities, possibly influenced by unrecognized pleiotropy.

Although the pleiotropy test showed that the *p*-value for the Egger intercept was 0.3973, suggesting that the influence of pleiotropy on the results might be limited, the findings should still be interpreted with caution. Additionally, the results from the Weighted Median method, Simple Mode method, and Weighted Mode method did not indicate a significant causal relationship. The OR for the Weighted Median method was 2.084*×*10^-6^ (95% CI: 2.49239*×*10^-21^–1742471179, *p* = 0.4555), and for the Simple Mode method, the OR was 6.81772*×*10^-6^ (95% CI: 1.8768*×*10^-27^–2.47663*×*10^16^, *p* = 0.6552) (Table 3). These results also displayed extremely wide confidence intervals, further indicating that the current evidence is insufficient to support a clear causal relationship between Selenium and congenital foot deformities. Furthermore, heterogeneity tests (Table 4) revealed no significant heterogeneity in either the IVW method or the MR Egger method (IVW method: Q = 3.504786, df = 6, *p* = 0.7433; MR Egger: Q = 2.648841, df = 5, *p* = 0.7539) (Table 5), indicating good consistency among the effects of different SNPs. Despite this, the pleiotropy test suggests that potential pleiotropy could still influence the results, as indicated by the Egger intercept *p*-value of 0.3973.

Although the analysis results suggest a potential association between Selenium and congenital foot deformities, the wide confidence intervals, lack of statistically significant p-values, and potential influence of pleiotropy indicate that the current evidence is insufficient to conclude a clear causal relationship between Selenium and congenital foot deformities. Further large-scale studies are needed to validate these preliminary findings and enhance the reliability of the conclusions.

### 2.3 The causal relationship between Copper and congenital foot deformities

The causal association between Copper as an exposure factor and congenital foot deformities was systematically evaluated using Mendelian randomization. Copper, as an essential trace element, is involved in various biological processes, including enzyme activity regulation, iron metabolism, oxidative stress response, and neural development. Although Copper plays a crucial role in human development, its relationship with congenital deformities, particularly foot deformities, has not been thoroughly studied.

Firstly, we assessed the causal relationship between Copper levels and congenital foot deformities using the Inverse Variance Weighted (IVW) method. The results showed that there was no significant causal association between Copper and congenital foot deformities (*b* = -0.03688, SE = 0.08030, *p* = 0.64609, OR = 0.9638, 95% CI: 0.8234–1.1281) (Tables 2 and 6). Although the confidence interval suggests that Copper might have a protective effect, the association did not reach statistical significance, indicating that the relationship between Copper and congenital foot deformities may not be strong or is subject to some uncertainty. In the MR Egger regression analysis (Table 3), the association between Copper and congenital foot deformities remained non-significant (*b* = -0.05475, SE = 0.11925, *p* = 0.65699, OR = 0.9467, 95% CI: 0.7494–1.1960). This result further supports the lack of a significant causal relationship between Copper and congenital foot deformities. However, the MR Egger analysis suggests that pleiotropy may have introduced some bias into the results, which, while not statistically significant, should be considered in future research.

Additionally, in the heterogeneity tests (Table 4), the Q test results for both the MR Egger and IVW methods did not show significant heterogeneity (MR Egger: Q = 12.46954, df = 9, p = 0.1881; IVW: Q = 12.532, df = 10, p = 0.2510). This indicates that the SNPs used in the analysis of the relationship between Copper and congenital foot deformities exhibited consistent overall effects, with no significant heterogeneity observed. These results suggest that the metabolic pathways of Copper in the body are complex and varied, possibly exerting a potential protective effect through indirect mechanisms rather than directly influencing the occurrence of congenital foot deformities. Furthermore, the potential protective effect of Copper may be modulated or diminished by other biological factors, a hypothesis that warrants further investigation in future studies.

### 2.4 The causal relationship between Iron (Fe) and congenital foot deformities

Using the Mendelian randomization method, we systematically explored the causal relationship between Iron (Fe) as an exposure factor and congenital foot deformities, conducting an in-depth analysis with our own data. Iron is a crucial trace element in the human body, extensively involved in various biological processes such as oxygen transport, DNA synthesis, and energy metabolism. However, both excessively high and low Iron levels can potentially have adverse effects on normal fetal development, making it essential to further understand the relationship between Iron and congenital deformities.

Firstly, the causal relationship between Iron levels and congenital foot deformities was evaluated using the Inverse Variance Weighted (IVW) method. The results indicated no significant causal association between Iron levels and congenital foot deformities (*b* = -3.7895, SE = 7.0854, *p* = 0.5928) (Table 2), with an OR of 0.0226 (95% CI: 2.104*×*10^-8^–24289.3153) (Table 6). Similarly, in the MR Egger regression analysis, the association between Iron and congenital foot deformities was not significant (*b* = 15.2767, SE = 23.3454, *p* = 0.5216), with an OR of 4310892.21 (95% CI: 5.788*×*10^-14^–3.211*×*10^26^) (Table 6). Although the OR in the MR Egger analysis was extremely high, suggesting a potential association, the extreme width of the confidence interval and the non-significant *p*-value indicate considerable uncertainty and potential pleiotropic bias. The heterogeneity analysis showed no significant heterogeneity in the association between Iron and congenital foot deformities across the SNPs, whether using the MR Egger or IVW method (MR Egger: Q = 9.9609, df = 17, *p* = 0.9052; IVW: Q = 10.6956, df = 18, *p* = 0.9068) (Tables 4 and 5). This suggests that the effects of the selected SNPs were consistent and not significantly influenced by heterogeneity.

Additionally, the pleiotropy test results indicated that the intercept of the MR Egger analysis (-0.0500, SE = 0.0584, p = 0.4033) did not reach significance, suggesting that pleiotropy might have a minimal impact on the study results, though its influence cannot be entirely ruled out. This finding further highlights the complexity of the relationship between Iron and congenital foot deformities; however, the current data do not provide strong evidence to support this association. Other Mendelian randomization methods, such as the Weighted Median method, Simple Mode method, and Weighted Mode method, yielded consistent results with the aforementioned analyses, none of which reached statistical significance.

Some analysis results suggest a possible association between Iron levels and congenital foot deformities, particularly in cases with extreme OR values. However, the wide confidence intervals and non-significant p-values indicate substantial uncertainty and potential bias in the study. The current evidence is insufficient to conclude a strong causal relationship between Iron and congenital foot deformities. Future research should focus on exploring the mechanisms by which Iron influences fetal development, which could help better understand its potential relationship with congenital deformities and provide a scientific basis for related prevention and treatment strategies.

### 2.5 The causal relationship between Phosphate and congenital foot deformities

The analysis of the association between Phosphate as an exposure factor and congenital foot deformities showed varying degrees of correlation among several SNPs. For example, SNPs such as rs10273103, rs12132412, rs12308036, and rs146992721 displayed different *β* values and *p*-values, reflecting the potential impact of Phosphate on congenital foot deformities. Based on the results of Mendelian randomization analysis, we can observe that although certain SNPs, such as rs146992721, exhibited relatively large effects (*β* = 0.171084), the overall statistical significance was not strong (*p* = 0.185737). This suggests that the influence of Phosphate on congenital foot deformities may not be significant.

In further analysis, various Mendelian randomization methods, including the Inverse Variance Weighted (IVW) method, MR Egger method, and Weighted Median method, were used to validate these findings. The overall effect size obtained from the IVW method (*β* = *−*0.04388, *p* = 0.811944) did not show significant statistical significance, indicating that, overall, Phosphate exposure may not have a significant causal relationship with congenital foot deformities. Additionally, the results from the MR Egger method and the Weighted Median method also supported this conclusion, with the MR Egger method yielding a *p*-value of 0.9509, further excluding the possibility of bias.

In the heterogeneity tests, the Q statistics yielded high p-values (MR Egger: P = 0.7023; IVW: P = 0.7881), indicating good consistency among the effects of the various SNPs, with no significant heterogeneity observed. For the pleiotropy test, the intercept from the MR Egger method was close to zero (intercept = 0.0026, P = 0.9853), suggesting that pleiotropic effects were minimal and unlikely to affect the overall causal inference. Although some SNPs showed a potential association between Phosphate and congenital foot deformities, the overall Mendelian randomization analysis results do not support a significant causal relationship between Phosphate and congenital foot deformities. These findings suggest that Phosphate may not be a major contributing factor to congenital foot deformities, and further research, potentially with larger sample sizes or alternative methods, may be needed to validate these preliminary findings.

### 2.6 The Impact of Maternal Selenium Supplementation on Gut Microbiota and Its Role in Congenital Foot Deformities

A systematic evaluation was conducted to assess the causal relationship between maternal selenium supplementation during pregnancy and congenital foot deformities, with a particular focus on the potential mediating role of the gut microbiota, specifically class Bacilli. Using multiple analytical methods (such as Inverse Variance Weighted (IVW) method, MR Egger method, and the Weighted Median method), the study found a significant association between selenium and the gut microbiota class Bacilli (Table 7). The IVW results showed a p-value of 1.0567 *×* 10*^−^*^10^, suggesting a strong regulatory effect of selenium on this bacterial class (Table 8). Although the sensitivity analysis did not find a statistically significant direct effect of selenium on congenital foot deformities, the indirect effect of selenium was significantly enhanced when class Bacilli was considered as a mediator, indicating that selenium might reduce the risk of congenital foot deformities indirectly through its regulation of the gut microbiota (Table 9). Further genetic analysis revealed that several SNPs associated with class Bacilli, such as rs2952183 and rs11837200, were significantly related to this bacterial class, with p-values of 0.00001, supporting the possibility that selenium may influence fetal developmental abnormalities through genetic variations regulating gut microbiota (Table S1). Additionally, in the Mendelian randomization analysis, while the IVW method did not show a significant association between class Bacilli and congenital foot deformities (p = 0.1316), the Weighted Median method demonstrated a significant correlation, with a p-value of 0.0063 (Table 9), further supporting the critical mediating role of gut microbiota in the relationship between selenium and congenital foot deformities. Although the overall results did not reach statistical significance, this study provides new insights into the potential regulatory mechanisms between maternal nutrition, gut microbiota, and fetal developmental abnormalities. These findings offer a new direction for future research into the effects of trace elements on fetal development, highlighting the key role that gut microbiota may play in this process.

**Table 7.**
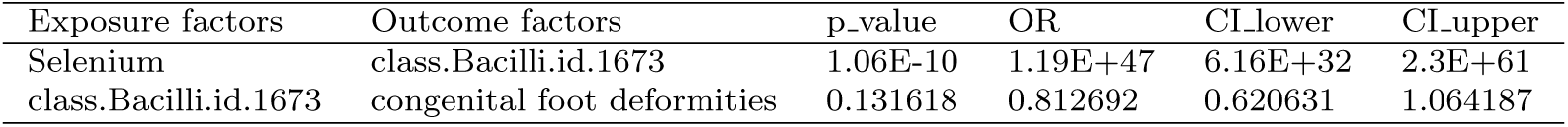
Gut Microbiota as a Mediator, Selenium, and Congenital Foot Deformities OR and Related Data.

**Table 8.** Gut Microbiota as a Mediator, Selenium, and Congenital Foot Deformities - IVW Method.

| Exposure factors | Outcome factors | method | nsnp | b | se | pval |
| --- | --- | --- | --- | --- | --- | --- |
| Selenium | class.Bacilli.id.1673 | Inverse variance weighted | 64 | 108.3969 | 16.78333 | 1.06E-10 |
| class.Bacilli.id.1673 | congenital foot deformities | Inverse variance weighted | 61 | -0.2074 | 0.137558 | 0.131618 |

**Table 9.**
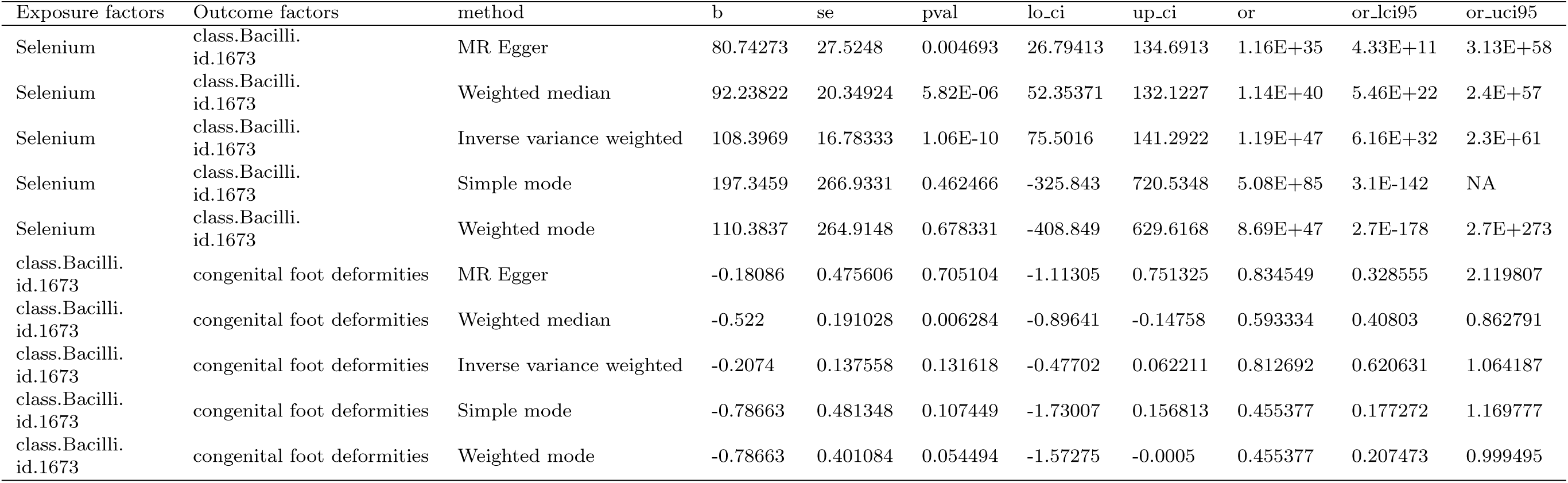
Gut Microbiota as a Mediator, Selenium, and Congenital Foot Deformities - Mendelian Randomization Using Five Method.

### 2.7 Visualization analysis

This study systematically explored the potential causal relationships between zinc, selenium, copper, iron, and phosphate with congenital foot deformities. Additionally, through visual analysis, it evaluated the effect of selenium on the gut microbiota, specifically class Bacilli, and its mediating role in congenital foot deformities. The analysis results showed that genetic variations in zinc significantly increased the risk of congenital foot deformities. Forest plots and IVW analysis (Figures 1 and 3) further supported the important role of zinc as a cofactor in various enzymes during fetal development, participating in key biological processes such as DNA synthesis, cell division, and protein synthesis. Notably, SNPs like rs1175550 and rs2769264 demonstrated significant effects in the analysis, indicating a significant association between zinc genetic variations and the risk of congenital foot deformities. Conversely, the associations of selenium, copper, iron, and phosphate with congenital foot deformities did not reach statistical significance in the leave-one-out and sensitivity analyses, although some SNPs, such as rs146992721, exhibited large effect sizes. However, the overall associations remained weak (Figures 2 and 4).In the analysis of selenium and gut microbiota, significant associations between selenium and Bacilli were observed using the IVW, MR Egger, and weighted median methods (Figure 6), with several SNPs, such as rs74663707 and rs12642660, showing significant effects on Bacilli. The leave-one-out analysis confirmed the robustness of this result, while the funnel plot indicated that the SNPs were symmetrically distributed around the central axis, suggesting no obvious bias (Figure 7).Overall, the results suggest that selenium may indirectly influence the risk of congenital foot deformities by regulating class Bacilli8.

**Fig. 1.**
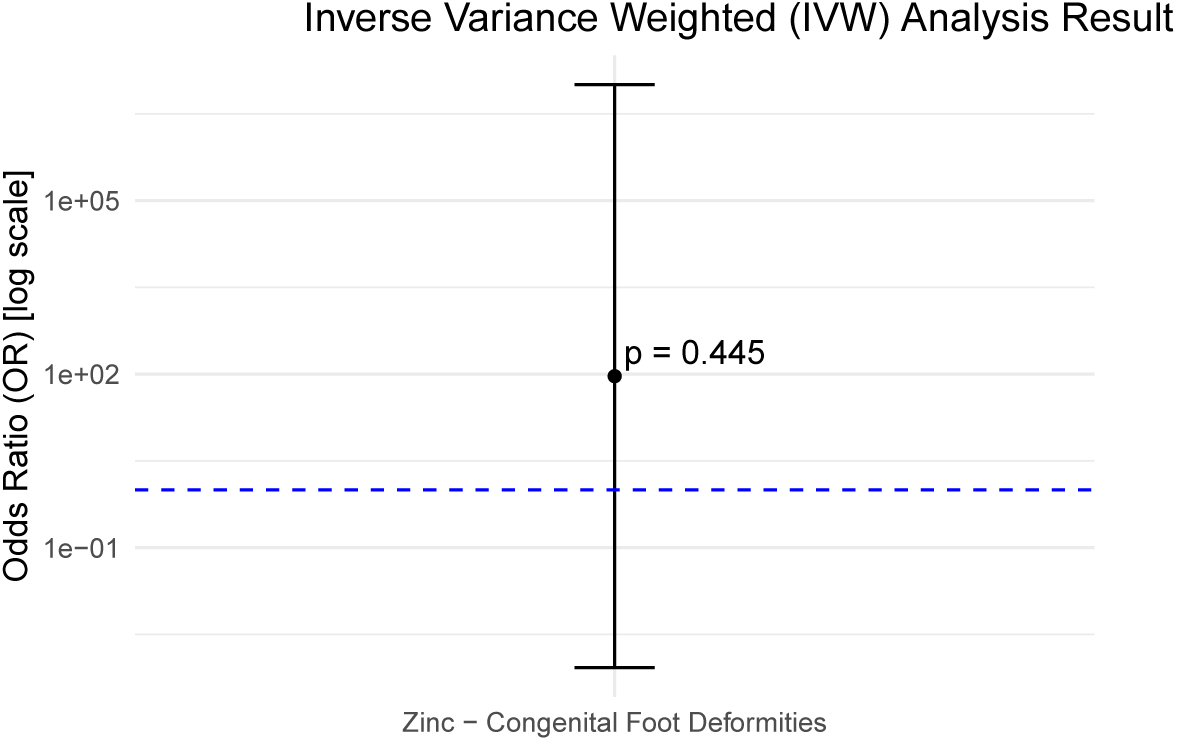
IVW Analysis Result of Zinc

**Fig. 2.**
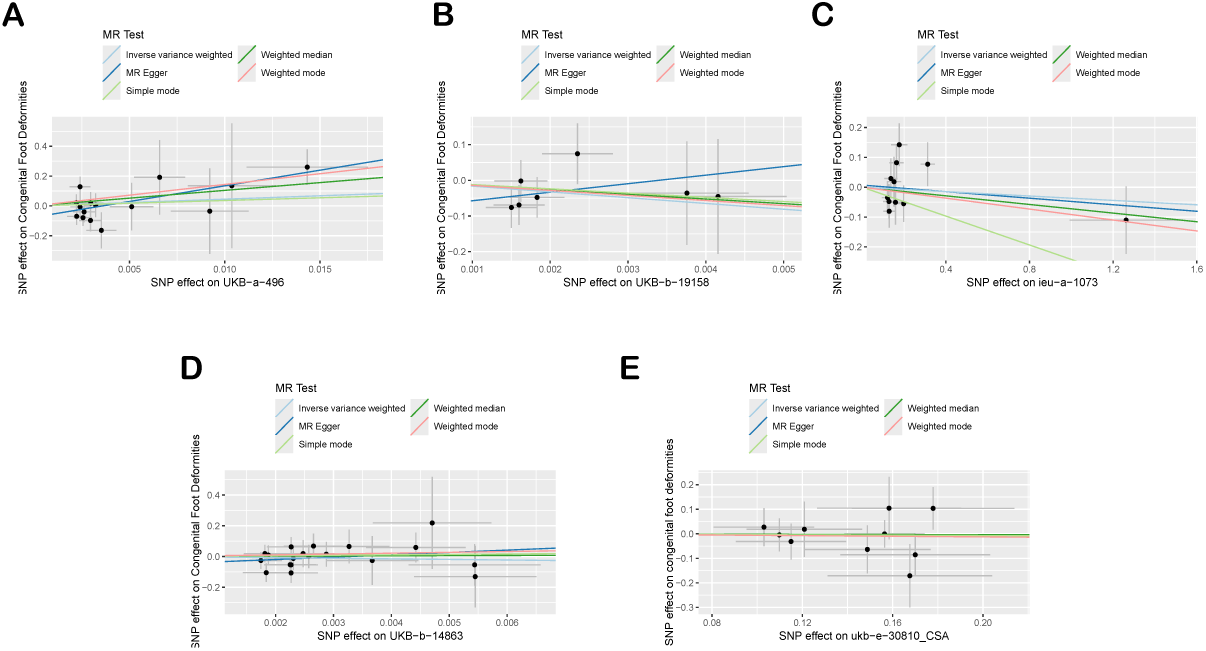
Scatter Plot

**Fig. 3.**
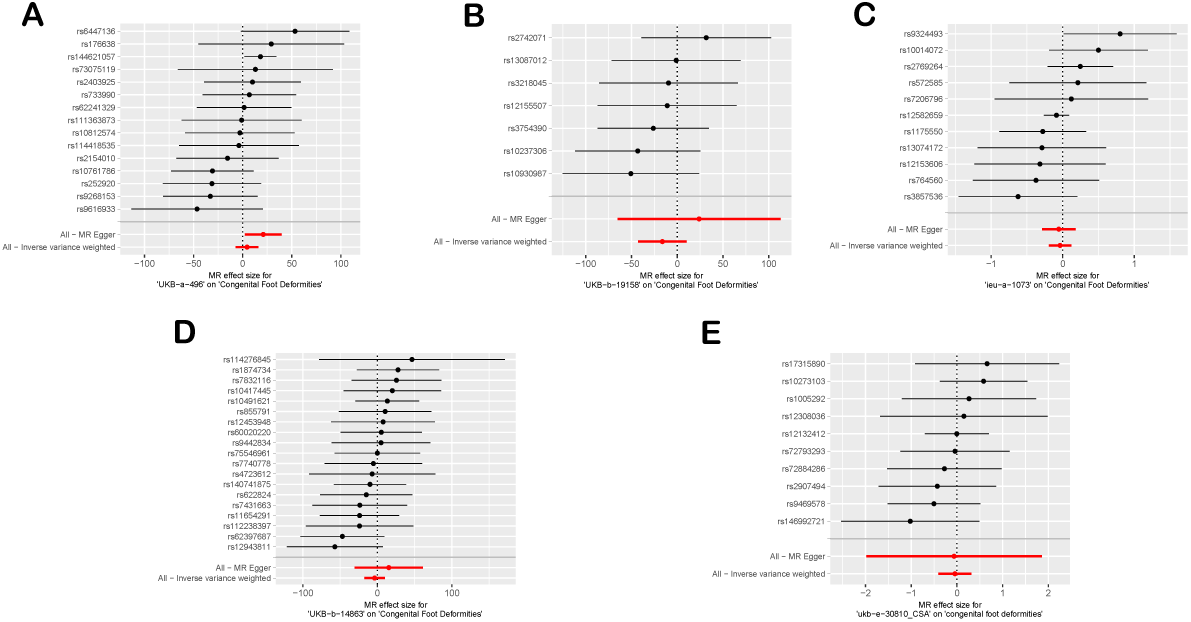
Forest Plot

**Fig. 4.**
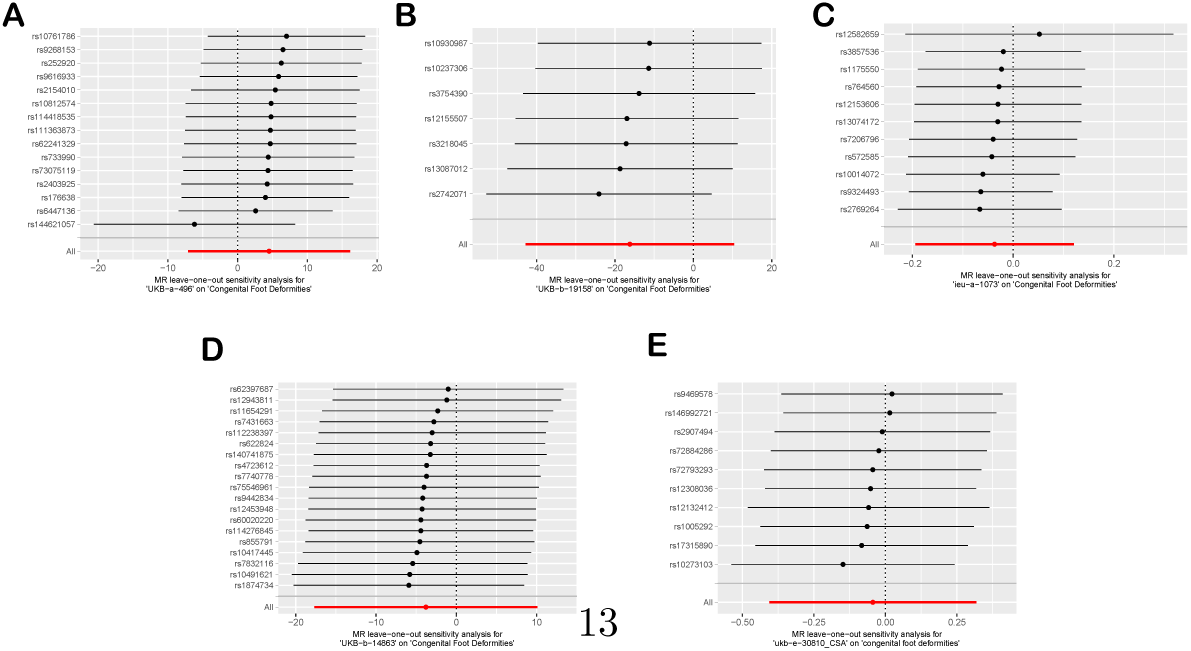
Leave-One-Out Plot

**Fig. 5.**
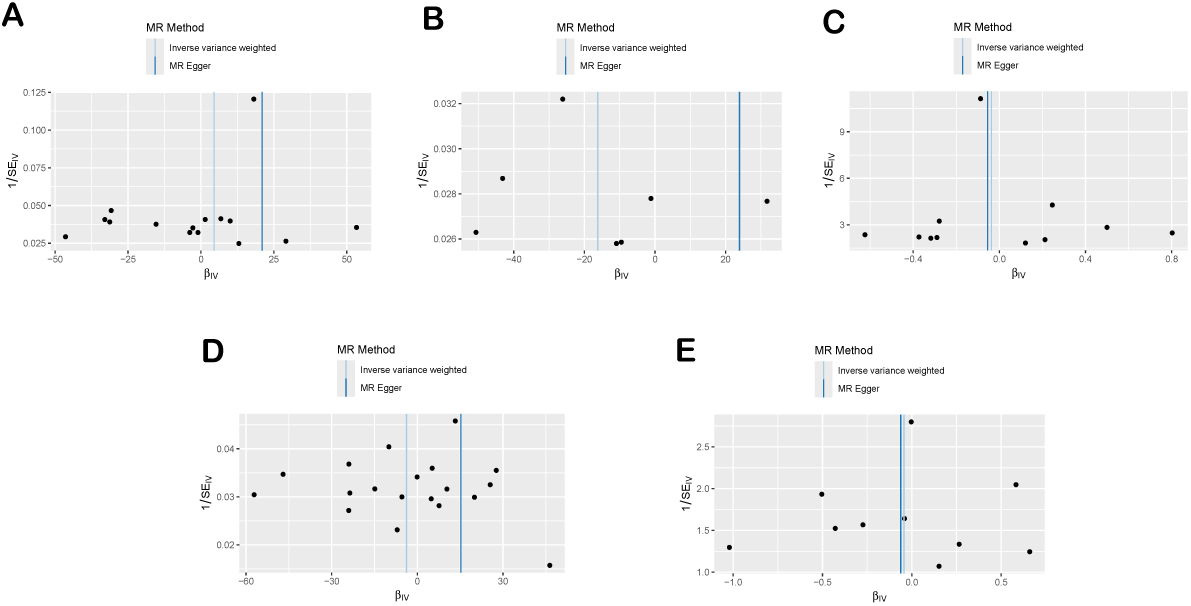
Funnel Plot

**Fig. 6.**
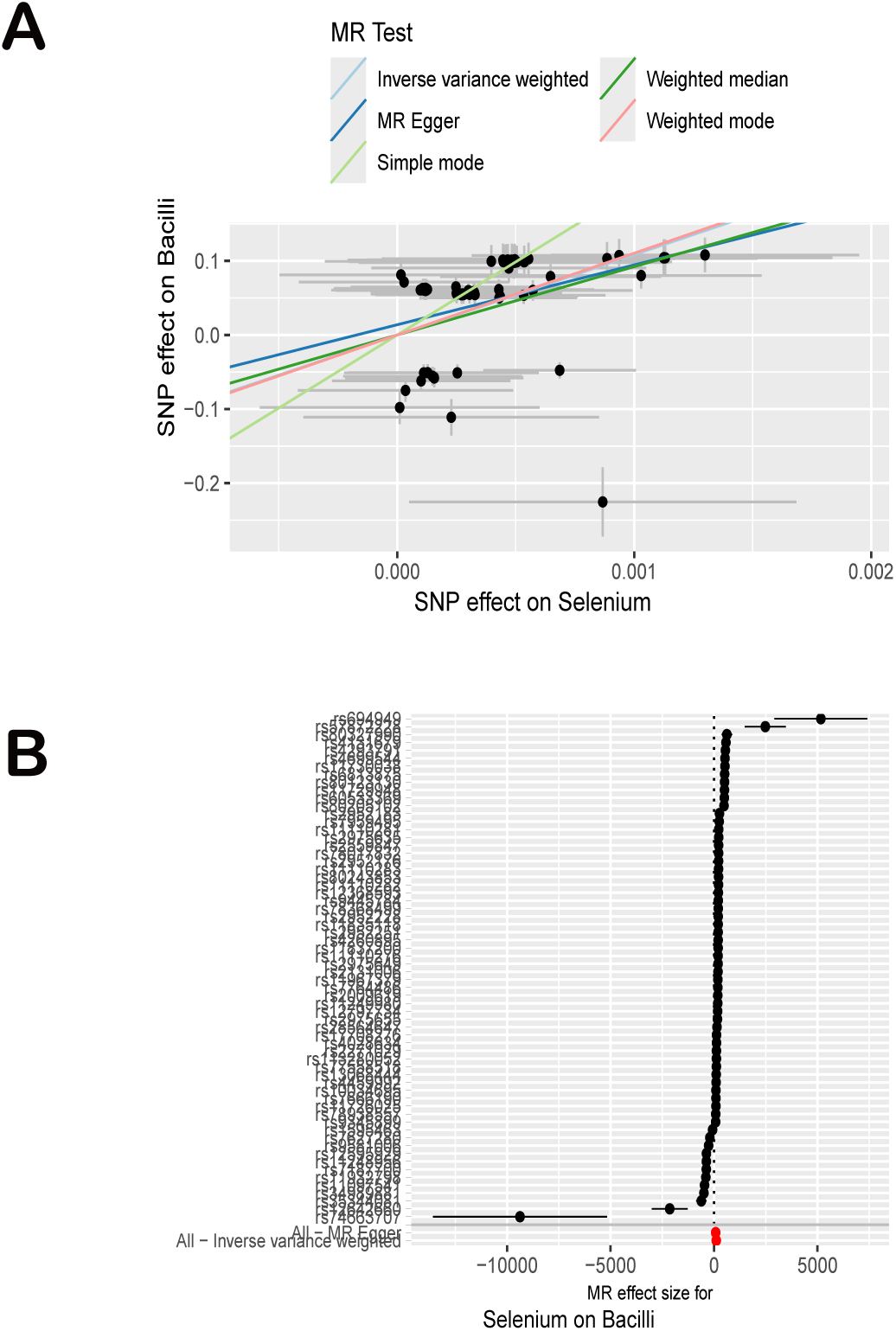
Scatter and forest plots of Selenium and Bacilli

**Fig. 7.**
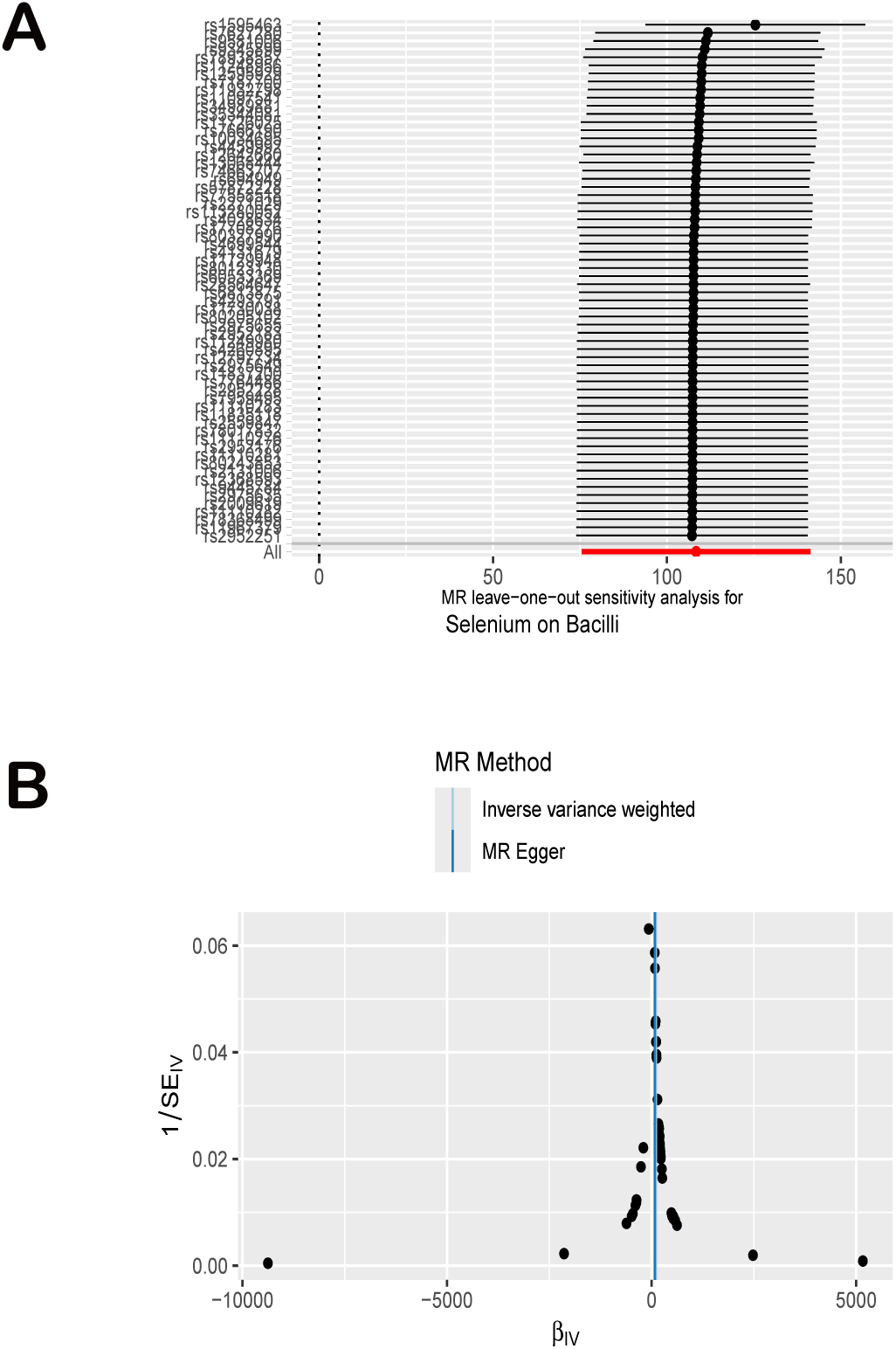
Leave-one-out and funnel plots of Selenium and Bacilli

**Fig. 8.**
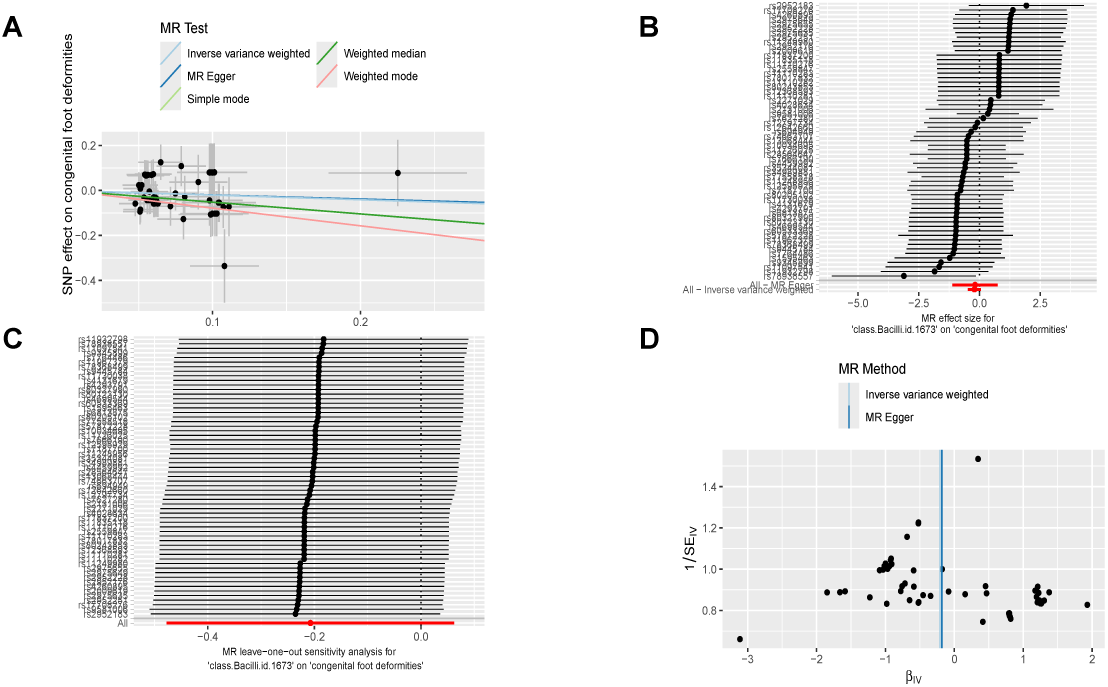
Randomization Visualization Analysis of Bacilli and Congenital Foot Deformities

## 3 Methods

### 3.1 Study Design

This study employed the Mendelian Randomization (MR) method to investigate the causal relationships between the levels of trace elements Zinc, Selenium, Copper, Iron, and Phosphate and the risk of congenital foot deformities. Mendelian Randomization is an analytical method that uses genetic variations as instrumental variables to infer causal relationships between exposure factors and outcomes, effectively avoiding the confounding factors and reverse causality issues common in traditional observational studies[27]. The analysis in this study was based on data from large-scale Genome-Wide Association Studies (GWAS)[28].

The exposure variables in this study were the genetically predicted levels of the trace elements Zinc, Selenium, Copper, Iron, and Phosphate. The exposure data were sourced from published summary statistics of Genome-Wide Association Studies (GWAS) conducted in populations of European ancestry. The gut microbiota data primarily comes from the MiBioGen Consortium’s GWAS summary data, focusing on SNPs that are significantly associated with the gut microbiota (*p ≤* 1*×*10*^−^*^5^).To ensure the robustness of the analysis, the genetic instruments included were single nucleotide polymorphisms (SNPs) significantly associated with Selenium, Iron, and Phosphate levels (*p ≤* 5 *×* 10^-6^), and with Zinc and Copper levels (*p ≤* 1 *×* 10^-5^). Linkage disequilibrium (LD) clumping (*r* ^2^ *<* 0.001, window size = 10,000 kb) was used to remove SNPs in LD. The causal relationship between trace element levels and congenital foot deformities was evaluated using the Inverse Variance Weighted (IVW) method. Sensitivity analyses and assessments of pleiotropy and heterogeneity were conducted using MR-Egger regression, the Weighted Median method, and Cochran’s Q test. All analyses were performed using the “TwoSampleMR” package in R software. Hypothesis testing was two-sided, with a significance level set at *p <* 0.05, and Bonferroni correction was applied, adjusting the significance level to *p <* 0.01. All results were reported with 95% confidence intervals (CI).

### 3.2 Main analysis method

The primary analysis method was the Inverse Variance Weighted (IVW) approach, which assumes that all instrumental variables are valid to provide optimal statistical power. Additionally, MR-Egger regression and the Weighted Median method were used for sensitivity analyses to detect potential pleiotropy and biases in the instrument variables’ validity. First, SNPs associated with the exposures were extracted from the GWAS summary data, and the effects of these SNPs on the outcomes were estimated. Then, the IVW method was used to combine the effect estimates of the SNPs to obtain an overall estimate of the causal effect. To further assess the robustness of the results, sensitivity analyses were conducted using MR-Egger regression and the Weighted Median method to check for pleiotropy and other potential biases in the instrumental variables.

### 3.3 Statistical analysis

By utilizing a combination of scatter plots, forest plots, leave-one-out plots, and funnel plots, we conducted a comprehensive graphical analysis to assess the robustness and consistency of the causal relationships between the trace elements Zinc, Selenium, Copper, Iron, and Phosphate and congenital foot deformities. The scatter plot illustrated the relationship between SNP effect estimates and their standard errors, providing a preliminary basis for understanding the effect distribution and identifying potential biases. The forest plot further visualized the effects of individual SNPs along with their 95% confidence intervals, helping to confirm the contribution and consistency of each SNP to the overall effect. The leave-one-out plot, which recalculates effect estimates by sequentially excluding each SNP, validated the robustness of the results and ruled out dependence on any single SNP. The funnel plot, by displaying the symmetry between effect estimates and standard errors, was used to detect potential publication bias and pleiotropy. These graphical analysis methods complemented each other, collectively providing a solid evidence base for the study, ensuring the reliability and accuracy of the results. **Ethical approval declarations** All the data used in this study were derived from publicly available GWAS studies, all of which had received approval from the respective ethics committees, and all participants had provided informed consent. Therefore, no additional ethical approval was required for this study. Through the methods outlined above, we aim to clarify the causal relationships between the trace elements Zinc, Selenium, Copper, Iron, and Phosphate and congenital foot deformities. This study seeks to provide new genetic evidence for understanding the role of these trace elements in fetal development and to offer a theoretical basis for nutritional management during pregnancy.

If your manuscript includes potentially identifying patient/participant information, or if it describes human transplantation research, or if it reports results of a clinical trial then additional information will be required. Please visit (https://www.nature.com/nature-research/editorial-policies) for Nature Port-folio journals, (https://www.springer.com/gp/authors-editors/journal-author/journal-author-helpdesk/publishing-ethics/14214) for Springer Nature journals, or (https://www.biomedcentral.com/getpublished/editorial-policies#ethics+and+consent) for BMC.

## 4 Discussion

In this study, we employed the Mendelian randomization method, utilizing genetic variants as instrumental variables to mitigate confounding biases and reverse causation issues commonly present in traditional observational studies. This approach allowed for a more accurate assessment of the impact of these trace elements on congenital foot deformities. Through Mendelian randomization analysis, we systematically evaluated the potential causal relationships between trace elements—zinc, selenium, copper, iron, and phosphate—and congenital foot deformities. We found a significant association between zinc levels and the risk of congenital foot deformities, while selenium, copper, iron, and phosphate did not show similar causal relationships. These findings provide new insights into the roles of these trace elements in fetal development.

The analysis results indicate a significant association between genetically elevated zinc levels and the risk of congenital foot deformities. This finding aligns with the conclusions drawn from a substantial body of existing literature, further reinforcing the critical role of zinc in fetal development[29]. As a cofactor for numerous enzymes and transcription factors, zinc plays an essential role in crucial biological processes such as cell division, DNA synthesis, and protein synthesis[30]. Zinc deficiency during pregnancy has been shown to be associated with various adverse pregnancy outcomes, including preterm birth, low birth weight, and developmental abnormalities[31]. This finding is consistent with the existing literature. For instance, Adams pointed out in their systematic review that zinc deficiency may lead to developmental disorders of fetal bones and soft tissues, thereby increasing the risk of congenital malformations[32]. Additionally, experimental studies have demonstrated that adequate zinc supply helps regulate signaling pathways and gene expression during fetal development, further supporting the findings of our study[33]. During the critical stages of fetal development, appropriate zinc levels are vital for cell differentiation and organ formation. If zinc levels are insufficient during this period, it may lead to cellular dysfunction, thereby affecting the normal embryonic development process and increasing the risk of congenital malformations[34]. Combining our data analysis, specific SNP loci such as rs1175550 and rs2769264 showed significant effect values between zinc exposure and the risk of congenital foot deformities (e.g., the *β* value for rs1175550 was 0.198, with a significant P value), suggesting that these genetic variants may influence embryonic development by regulating zinc levels in the body, thereby increasing the probability of congenital foot deformities. These results not only validate the importance of zinc in fetal development but also provide a scientific basis for further exploration of zinc’s potential role in preventing congenital malformations. Moreover, this finding suggests that monitoring and appropriately supplementing zinc levels during pregnancy could be an effective strategy to reduce the risk of congenital foot deformities[35].

Compared to the findings on zinc, this study did not identify significant causal relationships between selenium, copper, iron, and phosphate and congenital foot deformities. Selenium is believed to have antioxidant and immune-regulatory functions that can protect cells from oxidative stress damage[36]. However, despite some epidemiological studies reporting potential associations between selenium intake during pregnancy and the risk of birth defects[37], our Mendelian randomization analysis did not support this hypothesis. This may indicate that selenium has a minor impact on congenital foot deformities, or that other environmental factors play a more critical role. Similarly, copper and iron are essential trace elements in fetal development, involved in processes such as hematopoiesis and redox reactions[38]. Nevertheless, this study did not find significant causal relationships between these two elements and congenital foot deformities. This may be related to the metabolic complexity of copper and iron. For example, Huang and colleagues pointed out that although copper deficiency is associated with neural tube defects, its impact on other types of congenital malformations remains unclear[39]. Additionally, iron intake and utilization are regulated by various factors, such as maternal iron metabolism and placental function, which may influence its specific role in fetal development[40]. Regarding phosphate, although its role in skeletal development is undeniable, our analysis did not find a direct causal relationship between phosphate and congenital foot deformities. A possible explanation is that the fetus can typically obtain sufficient phosphate through maternal regulatory mechanisms, so changes in phosphate levels are unlikely to significantly affect foot development unless under extreme conditions such as severe malnutrition or metabolic disorders[41].

The findings of this study have significant public health and clinical implications. First, the significant association between zinc levels and congenital foot deformities highlights the importance of zinc supplementation during pregnancy[42] . Pregnant women should, under the guidance of a physician, consume zinc-rich foods or supplements to maintain adequate zinc levels, which may help reduce the risk of fetal congenital malformations. This finding provides a basis for public health policymakers to recommend increased attention to zinc levels in prenatal nutrition monitoring[43]. On the other hand, although selenium, copper, iron, and phosphate are undeniably important in fetal development, our study did not find significant causal relationships between these trace elements and congenital foot deformities. This result may reflect that the role of these elements in the occurrence of congenital malformations is more indirect or regulated by other factors. Therefore, clinically, there may be a greater focus on the impact of these elements on other health outcomes rather than on specific congenital malformations.

A Mendelian randomization analysis explored the relationship between selenium, the gut microbiota class Bacilli, and congenital foot deformities. The results showed a significant association between selenium and class Bacilli, suggesting that selenium may indirectly reduce the risk of congenital foot deformities by regulating the gut microbiota. Although the direct causal relationship between selenium and congenital foot deformities did not reach statistical significance, when Bacilli was considered a mediator, the indirect effect of selenium was significantly enhanced, indicating that selenium may exert its effects through the gut microbiota. This finding aligns with the conclusion of Rayman et al. [44], which suggests that selenium reduces fetal oxidative stress damage through its antioxidant properties. It is also consistent with Zeng et al.[45], who studied selenium’s role in modulating immune responses, further explaining selenium’s regulatory effect on class Bacilli. However, this study is in line with the view of Lee et al.[46], which suggests that multiple factors influence congenital deformities, indicating that selenium alone may not directly affect deformity formation but rather works through a complex mediating mechanism. Additionally, the important role of Bacilli in maintaining gut balance and regulating immune responses, as found by Willing et al.[47], further suggests that selenium may indirectly influence fetal development through the regulation of the gut microbiota.

## 5 Conclusion

This study used Mendelian randomization analysis to explore the causal relationships between trace elements—zinc, selenium, copper, iron, and phosphate—and congenital foot deformities. The results showed that genetically elevated zinc levels significantly increased the risk of congenital foot deformities, suggesting that appropriate zinc intake during pregnancy is crucial for preventing such malformations. In contrast, selenium, copper, iron, and phosphate levels were not significantly associated with congenital foot deformities, indicating that the effects of these trace elements may be more indirect or influenced by other factors. However, when Bacilli acted as a mediator, the indirect effect of selenium was significantly enhanced. A significant association was observed between selenium and the gut microbiota class Bacilli, suggesting that selenium may reduce the risk of congenital foot deformities indirectly by regulating the gut microbiota. This study provides new evidence for the critical role of zinc in fetal development and the potential protective role of selenium through the regulation of the gut microbiota class Bacilli in fetal development. It also highlights the need for future research to further explore the complex relationships between other trace elements, gut microbiota, and congenital malformations, providing a scientific basis for optimizing nutritional management during pregnancy and reducing the incidence of congenital malformations.

## Data Availability

All relevant data are within the manuscript and its Supporting Information files

## Supplementary information

If your article has accompanying supplementary file/s please state so here.

Authors reporting data from electrophoretic gels and blots should supply the full unprocessed scans for key as part of their Supplementary information. This may be requested by the editorial team/s if it is missing.

Please refer to Journal-level guidance for any specific requirements.

## Acknowledgements

This research was conducted at the Department of Orthopaedics, Jiujiang Hospital of Traditional Chinese Medicine, Jiujiang, Jiangxi. We would like to express our heartfelt gratitude to Ling Long for providing the financial support that made this study possible. We also extend our sincere thanks to all the authors for their valuable contributions throughout the entire research process, from study design and data analysis to manuscript preparation and revision.

## Declarations

Some journals require declarations to be submitted in a standardised format. Please check the Instructions for Authors of the journal to which you are submitting to see if you need to complete this section. If yes, your manuscript must contain the following sections under the heading ‘Declarations’:

- Funding This study was funded by Clinical Application and Research of Digital Technology in the Diagnosis and Treatment of Flatfoot in Children (Grant no. 20212BAG70004).
- Conflict of interest/Competing interests The authors declare no competing interests.
- Ethics approval and consent to participate The data used in this study were obtained from publicly available Genome-Wide Association Studies (GWAS) datasets, which have been ethically approved by the relevant institutions, and informed consent was obtained from the participants. Therefore, no additional ethical approval is required for this study.
- Consent for publication
- Data availability
- Materials availability
- Code availability
- Author contribution

L.L. and J.G. conceived the study and performed the primary data analysis. H.Y. contributed to the methodology and interpretation of results. G.D. supervised the project and provided critical revisions to the manuscript. X.X. and W.W. assisted in data collection and provided essential resources. L.L. and J.G. drafted the manuscript, and all authors critically reviewed and edited the final version. G.D. was responsible for the correspondence and project administration. L.L. provided the funding for this research. All authors have read and approved the final version of the manuscript. L.L. and J.G. contributed equally to this work.

If any of the sections are not relevant to your manuscript, please include the heading and write ‘Not applicable’ for that section.

Editorial Policies for:

Springer journals and proceedings: https://www.springer.com/gp/editorial-policies Nature Portfolio journals: https://www.nature.com/nature-research/editorial-policies *Scientific Reports*: https://www.nature.com/srep/journal-policies/editorial-policies BMC journals: https://www.biomedcentral.com/getpublished/editorial-policies

## Notes

### Competing Interest Statement

The authors have declared that no competing interests exist.

### Author Declarations

The data used in this study were obtained from publicly available Genome-Wide Association Studies (GWAS) datasets, which have been ethically approved by the relevant institutions, and informed consent was obtained from the participants. Therefore, no additional ethical approval is required for this study.

